# Mechanical Unloading is Associated with Decreased DNA Content in Cardiomyocytes Independent of Nucleation State

**DOI:** 10.1101/2020.12.18.20248510

**Authors:** Jun Luo, Stephen D. Farris, Deri Helterline, April Stempien-Otero

**Author notes:** Corresponding author: April Stempien-Otero, M.D. Box 358050, 850 Republican Street, Seattle, WA 98109, Email address. Luo Cardiomyocyte Remodeling after Mechanical Unloading.

## Abstract

**Background:** Cardiomyocytes increase DNA content in response to stress in humans. Proliferation has been reported in cardiomyocytes in failing hearts and following LVAD unloading which may represent a resolution of this process through cell division. However, cardiac recovery from LVAD is rare.

**Methods:** We quantified cardiomyocyte nuclear number, cell size, DNA content and the frequency of cell cycling markers by imaging flow cytometry from human subjects undergoing LVAD implantation or primary transplantation.

**Results:** Cardiomyocyte size was 15 percent smaller in unloaded versus loaded samples without differences in the percentage of mono-, bi, or multi-nuclear cells. DNA content per nucleus was significantly decreased in unloaded hearts versus loaded controls. Cell cycle markers, Ki67 and phosphohistone H3 (H3P) were not increased in unloaded versus failing samples.

**Conclusions:** Unloading of failing hearts is associated with decreased DNA content of nuclei independent of nucleation state within the cell. As these changes were associated with a trend to decreased cell size but not increased cell cycle markers, they may represent a regression of hypertrophic nuclear remodeling and not proliferation.

## INTRODUCTION

In mammals, cardiomyocyte (CM) proliferation is robust *in utero* but drops dramatically following birth ^1, 2^. To allow further growth of the heart, CMs undergo cellular hypertrophy and engage in cell cycle activity resulting in multi-nucleation and increased chromosome copy number (ploidy) ^1, 3, 4^, process that is essentially complete by adulthood ^5^. However, in response to stress, cell cycle activity appears to be re-initiated ^6, 7^: hypertrophied human hearts have increased CM ploidy compared to age-matched controls ^8^. Because increases in ploidy appear to be proportionate to increased cardiac mass, it has been hypothesized that this state is adaptive to provide increased templates for the transcription and translation of proteins necessary for hypertrophy ^9^. Why cardiomyocytes fail to proliferate in response to stress is incompletely understood.

Study of the mechanisms of cardiomyocyte nuclear remodeling is challenging as mouse models are limited by significant differences in cardiomyocyte development and responses to stress. At baseline, mice have a higher incidence of multi-nucleation [80% vs 20% binuclear CM in humans ^9, 10^] and do not robustly increase ploidy in response to stressors such as pressure overload or myocardial infarction ^9^. Until recently, human studies have been limited to autopsy series ^11 1, 12^ as endomyocardial biopsy has significant risk and low tissue yield.

The growing use of LVAD implantation as a bridge to transplant has provided a potential model to understand this process. LVADs unload the failing left ventricle, thereby reducing mechanical stress on the heart. Patients have increased systemic perfusion, decreased neurohormonal stimulation, and may exhibit complete myocardial recovery albeit at a low rate ^13^. Preliminary studies have observed changes in DNA content from hearts following LVAD implantation, including increases in the proportion of diploid CM ^14^ and markers of cell cycle activity ^15^. Some authors have interpreted these data as evidence that proliferation increases with unloading. ^16^. However, most studies used histologic analyses in which definitive identification of cell borders in challenging. Moreover, all studies interrogated relatively small numbers of cells and the documentation of adequate ventricular unloading of subjects was not reported.

To more precisely define changes in nuclear structure with unloading and provide a future platform to understand mechanisms driving or inhibiting cardiomyocyte proliferation *<u>in humans</u>*, we used a novel technique, imaging flow cytometry. This platform, (branded ImageStream) allows precise identification of cardiomyocytes and quantification of nuclear state and DNA content within these cells. Furthermore, this strategy is high throughput facilitating analysis of ten times more cells per subject than previous studies. We isolated cardiomyocytes from subjects with advanced heart disease undergoing primary transplant or subjects who underwent unloading with LVAD prior to transplant. Unloading was confirmed by structural, hemodynamic and neurohormonal measures. Our data confirm previously reported nuclear changes, but suggest that these findings represent a more global process of nuclear re-organization occurring in the absence of increased cardiomyocyte cell cycle activity.

## METHODS

### Study Population and Design

Twenty-five subjects with end-stage systolic heart failure (20 males and 5 females; a majority with idiopathic cardiomyopathy) referred for heart transplantation at the University of Washington Medical Center were enrolled. Fifteen subjects underwent LVAD implantation for end-stage heart failure as a bridge to transplantation (CHF-Unloaded). Eight patients receiving primary heart transplantation without LVAD placement served as controls (CHF-Loaded). The seven subjects with ischemic cardiomyopathy were evenly distributed between the groups. Tissue from these seven subjects did not have evidence of infarction or scar on histologic sections adjacent to tissue that was processed for cell isolation. Two subjects with normal ventricular size and function who underwent transplant for coronary artery disease with chronic chest pain were also enrolled as controls (Control). Details on clinical data acquisition can be found in the Supplemental Methods.

### Isolation of Cardiac Cells

Cardiomyocytes were dissociated from fresh cardiac samples as detailed in the Supplemental Methods. To validate that our dissociation protocol yielded intact cardiomyocytes, cells were stained with antibodies against α-actinin, cardiac Troponin T, and pan-cadherin. Dissociated cardiomyocytes were smeared on a cover slip and imaged with fluorescent microscopy for tight junctions.

### Fluorescent Immunohistochemistry Labeling

Triple-fluorescent immunological staining was carried out on each sample. For identification of CM we used antibodies against sarcomeric α-actinin 2 µg/ml (clone EA-53, sigma A7723) Cellular DNA was stoichiometrically labeled by 10 µM Draq5 (ThermoFisher 62251) in flow cytometry buffer (100mM Tris-HCL, 300mM NaCl, 0.5mM MgCl_2_, 0.25mM CaCl_2_ 0.1% Triton-100). For cell cycling markers, rabbit anti-human Ki67 2 µg/ml (Abcam ab15580) or phosphorylated histone H3 2 µg/ml (H3P, Ser10, EMDMillipore 06-570) were used. Parallel staining procedures were carried out in cultured cardiac fibroblasts (isolation and culture detailed in the Supplemental Methods) with anti-human vimentin (RV202, Abcam ab8978).

### Imaging Flow Cytometry Analysis

To evaluate cardiomyocyte size, DNA content and frequency of positive proliferation markers of individual cardiomyocytes, immunofluorescently labeled cells were analyzed on an Amnis^®^ cellular imaging flow cytometer (ImageStream Mark II, EMD Millipore). Detailed methods can be found in Supplement and representative images are shown in Supplemental Figure 1.

### Statistics

Values are presented as mean ± one standard deviation. Unpaired student’s *t*-tests with Bonferroni correction were used for the comparison between two groups and ANOVA for comparing multiple groups unless noted otherwise. Simple linear regression was applied to analyze relationships, and coefficients demonstrating p-values less than 0.05 were considered significant.

### Study Approval

Informed consent was obtained from all subjects and the study protocols were approved by the UW Institutional Review Board.

## RESULTS

### Subject Characteristics

Tissue was collected from 8 subjects who underwent primary cardiac transplant (Loaded), 15 subjects bridged to transplant with LVAD (Unloaded), and two control subjects. Clinical data are in Table 1. All hearts in the Loaded and Unloaded groups had cardiomegaly and significant systolic dysfunction. Control subjects (one male, one female) had normal ejection fraction, left ventricular size and wall thickness. Subjects in the Unloaded group had significant regression of markers of hypertrophy at the time of transplant compared to pre-LVAD values. There was a 25% decrease in indexed cardiac mass per individual subject (106 ± 32 g/m^2^ post-LVAD versus 141 ± 33 g/m^2^ pre-LVAD, *P* = 0.002); smaller LVEDD (7.7 ± 1.0cm pre-LVAD vs 5.9 ± 0.9cm post-LVAD, *P* < 0.001) and 3-fold decrease in BNP (Supplemental Table 1).

**Table 1:** General Subject Characteristics. Data are presented as mean ± Standard deviation, * P < 0.05 vs. Loaded

|  | Normal EF | CHF-Loaded | CHF-Unloaded |
| --- | --- | --- | --- |
| N (male:female) | 2 (1:1) | 8 (5:3) | 15 (14:1) |
| Age (years) | 55.5 ± 12.5 | 51.7 ± 13.3 | 58.4 ± 9.1 |
| Heart Weight (HW, g) | 338 ± 71 | 529.5 ± 189.6 | 528.5 ± 99.9 |
| Body Weight (BW, kg) | 89.4 ± 10 | 77 ± 16.8 | 92.4 ± 14.8* |
| HW-BW Ratio (g/kg) | 3.7 ± 0.4 | 6.4 ± 2 | 5.8 ± 1.2 |
| EF (%) | 60 ± 8.5 | 21.4 ± 4.4 | 17.5 ± 4.2 |
| LVEDD (cm) | 5.0 ± 0.14 | 6.5 ± 1.6 | 5.8 ± 1.2 |
| Serum BNP (g/ml) | - | 1.1 ± 0.8 | 0.3 ± 0.3* |
| Duration of LVAD (days) | - | - | 387.9 ± 239.7 |

### Cardiomyocyte Morphologic Characteristics

Cardiomyocyte cell size, nuclear number and nuclear size were quantified using imaging cytometry on isolated cardiomyocytes. To validate that dispersed cardiomyocytes were intact, immunofluorescent staining of cells was performed with a pan-cadherin marker. Although the smear demonstrated the presence of free nuclei and cardiomyocyte fragments without nuclei (Figure 1A), all dispersed cardiomyocytes with intracellular nuclei had preserved sarcomeric structure (Figure 1B) and intact tight junction structures Figure 1B, C). Algorithms for image analysis also defined cardiomyocytes as having an aspect ratio (width/height) of less than 1.0 in addition to presence of α-actinin stain and intact nuclei (Supplemental Figure 1). There was a nearly significant trend toward decreased cell area (3019 ± 686 µm^2^ vs. 2574 ± 373.3 µm^2^, *P* = 0.06, Figure 2B) in Loaded versus Unloaded hearts. The duration of LVAD unloading had no effect on cell size (Figure 2C). CM size correlated strongly with total nuclear area in failing hearts (R^2^ = 0.3, *P* = 0.0037, Figure 2D). CM from the two control subjects had considerably smaller cell and nuclear area than both Loaded and Unloaded hearts (Figure 2D). To determine if these results were biased by the significantly higher body mass in the Unloaded group, we correlated cardiomyocyte size to heart mass for both groups. Cardiomyocyte size did not correlate to heart mass in either loaded or unloaded hearts (Supplemental Figure 2, R^2^ = 0.03 for each state).

**Figure 1:**
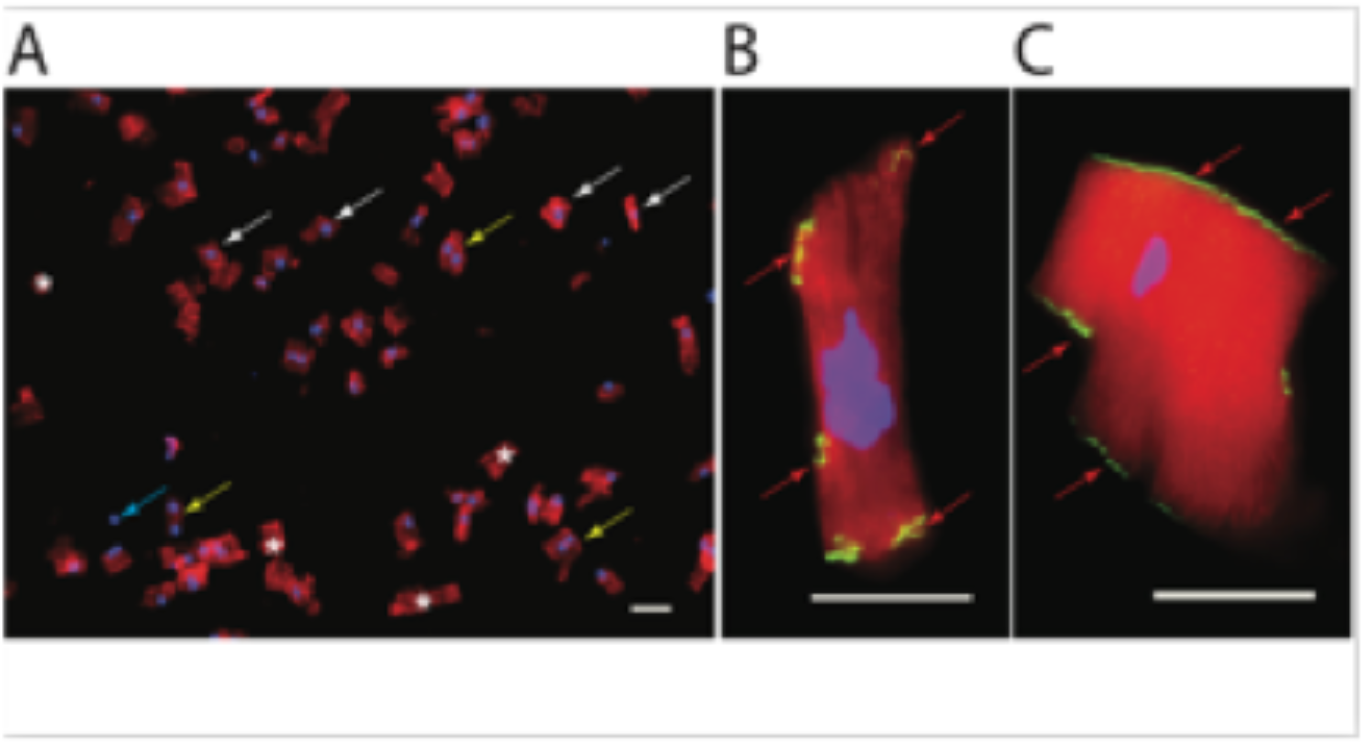
Validation of intact dispersed cardiomyocytes with immunocytochemistry. **A**. Representative image from smear of dissociated cardiomyocyte suspension prior to the imaging flow cytometry analysis. Immunofluorescence staining for α-actinin (red) and nuclei (blue), shows heterogeneous particles including dispersed mono-nuclear (white arrows), bi-nuclear cardiomyocytes (yellow arrows), and free nuclei (blue arrow) and myocyte fragments without nuclear content (stars). Dispersed cardiomyocytes with nuclei have preserved sarcomeric structure (**B**. red = α-actinin and **C**. red = cardiac Troponin T). In **B**. and **C**. dual labeling with antibody against pan-cadherin (green) demonstrates intact tight junction structures. Scale bar = 50 µm

**Figure 2.**
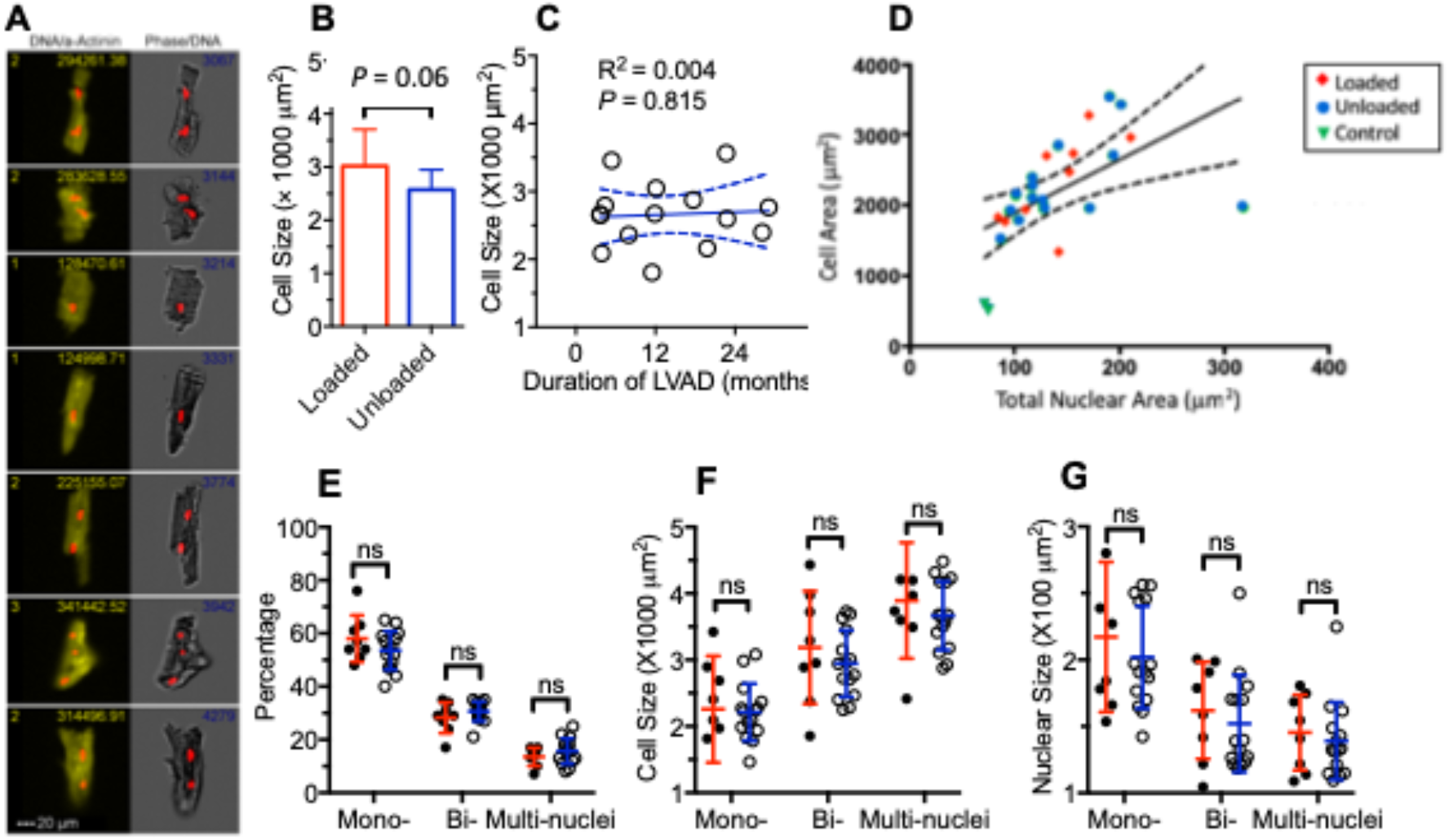
Effect of mechanical circulatory unloading on cardiomyocyte nucleation state, cellular and nuclear sizes. **A.** Representative images of cardiomyocyte during the imaging flow cytometry. **B.** Cardiomyocyte size from loaded and unloaded hearts. **C.** Relationship of cardiomyocyte size to duration of circulation support. **D.** Relationship between cardiomyocyte cell size and total nuclear area. **E.** Nucleation state from loaded and unloaded hearts. **F.** Average cardiomyocyte sizes at different nucleation level. **G.** Average nuclear sizes at different nucleation level. All data are presented as mean values ± standard deviation. Unpaired student’s *t*-tests with Bonferroni correction were used for the comparison between two groups. Simple linear regression was used in the correlation analysis (C and D). Each closed dot (loaded) or opened dot (unloaded) represent the mean value of one individual heart. Red lines represent loaded group, blue lines are unloaded group. Dotted line denoted 95% confidence band of the best-fit lines. P < 0.05 as statistic significant. ns = not significant.

Overall percentages of mono-bi- and multi-nucleated cells did not differ between CM from loaded versus unloaded hearts (mononucleated cells: 58% ± 9% vs. 53% ± 7%, *P* = 0.50; binucleated cells: 28% ± 6% vs. 30% ± 4%, *P* = 0.57; multinucleated cells: 14% ± 3% vs. 16% ± 5%, *P* = 0.62; Figure 2F). There were no differences in CM cell size between loaded and unloaded hearts at any nucleation state. However, cell size increased with multinucleation in both groups (mononucleated cells: 2258 ± 802 µm^2^ vs. 2212 ± 429 µm^2^, *P* = 1.00; binucleated cells: 3186 ± 849 vs. 2944 ± 502 µm^2^, *P* = 0.78; multinucleated cells: 3893 ± 873 vs. 3666 ± 517, *P* = 0.82; Figure 2G). Likewise, nuclear size did not differ with loading state but decreased with increasing nucleation state (mononucleated cells: 217 ± 56 µm^2^ vs. 202 ± 38 µm^2^, *P* = 0.83; binucleated cells: 162 ± 36 vs. 152 ± 37 µm^2^, *P* = 0.90; multinucleated cells: 146 ± 28 vs. 139 ± 29, *P* = 0.94; Figure 1H). ANOVA confirmed significant relationships between cell and nuclear size and nucleation state (*P* < 0.001 for both) but not unloading state.

### Cellular and Nuclear DNA Content is Reduced in Unloaded Hearts

Average DNA content per cardiomyocyte, as measured by the absolute fluorescence intensity in stoichiometrically labeled nuclei, was significantly reduced when comparing unloaded to loaded hearts (loaded: 157 ± 113 vs. unloaded: 72 ± 21, *P* = 0.009). Total DNA content of cardiomyocytes from the two hearts with normal ejection fraction was lower than both loaded and unloaded samples. To test the hypothesis that lower DNA content cardiomyocytes represented generation of new mononuclear cells, we measured DNA content by nucleation status. DNA content was significantly higher in loaded hearts from mono- (102 ± 70 vs. 52 ± 22, P = 0.046) and bi-nucleated (198 ± 133 vs. 99 ± 39, P = 0.039) but not multi-nucleated cardiomyocytes (205 ± 150 vs. 120 ± 50, P = 0.165; Figure 3A). Furthermore, the average DNA content <u>per nucleus</u> was decreased with unloading (mono-: 102 ± 70 vs. 52 ± 22, P = 0.046; bi-: 61 ± 40 vs. 30 ± 12, *P* = 0.03; multi-: 45 ± 34 vs. 24 ± 10, *P* = 0.113; Figure 3B). Moreover, although increased total cellular DNA content was strongly associated with cardiomyocyte size in hearts without LVAD use, this association was lost in the unloaded group (slope comparison: *P* < 0.0001 vs loaded subjects, Figure 3C). There was no association with the length of mechanical circulatory support on DNA content per cell (R^2^ = 0.020, *P* = 0.619, Figure 3D) or DNA content per nucleus (R^2^ = 0.0001, *P* = 0.971; Figure 3E).

**Figure 3.**
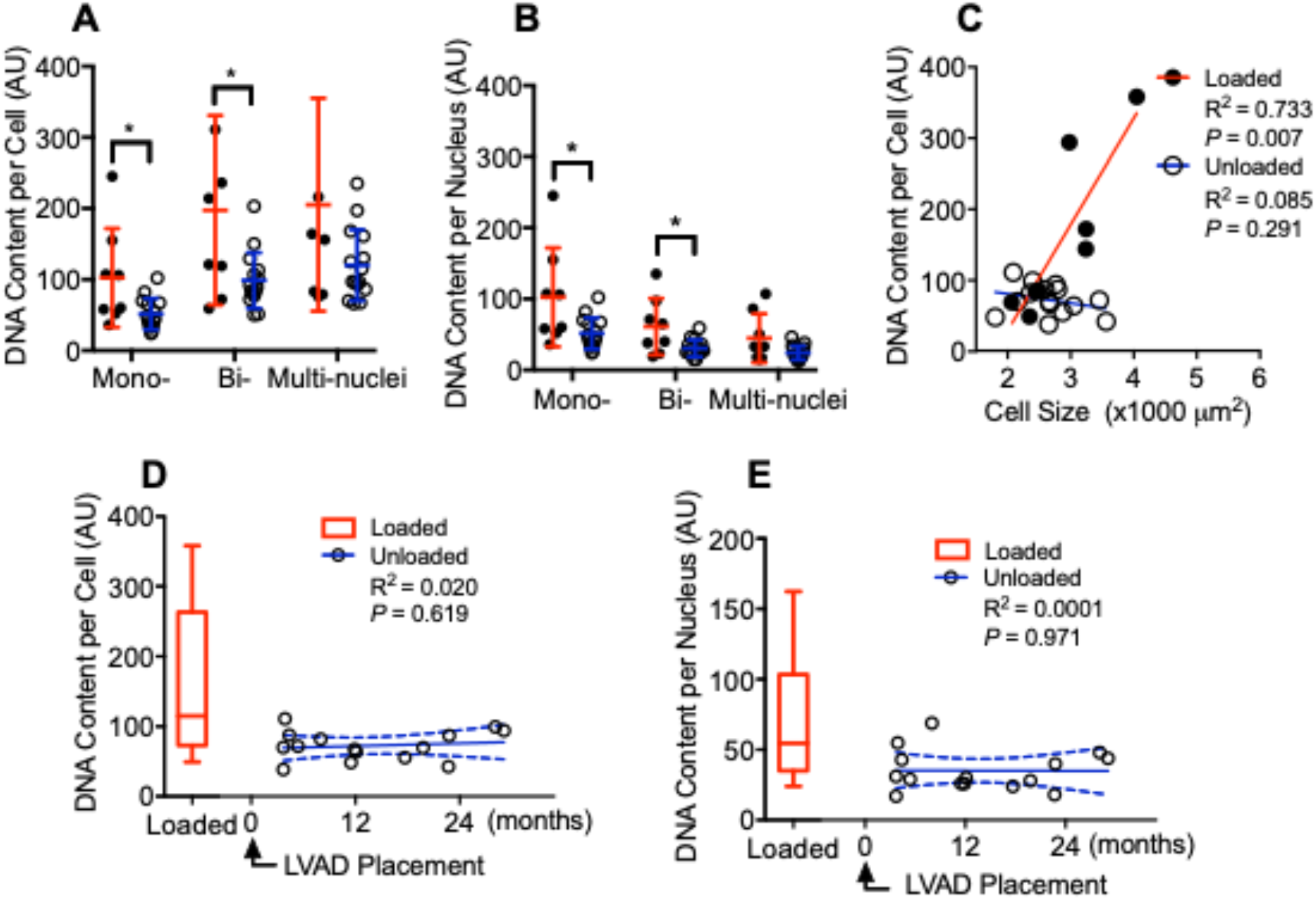
DNA content changes after mechanical circulatory unloading. **A**, DNA content per cardiomyocyte at different nucleation levels. **B**, DNA content per nucleus of each cardiomyocyte at different nucleation level. **C**, correlation of DNA content per cell and the average size of cardiomyocyte; **D**, correlation of DNA content per cardiomyocyte and the duration of circulatory unloading. **E**, correlation of DNA content per nucleus and the duration of circulatory unloading. All data are presented as mean values ± standard deviation. Unpaired student t test was used to compare the two groups (**A, B**). Simple linear regression was used in the correlation analysis (**C** - **E**). Each closed dot (loaded) or opened dot (unloaded) represent the mean value of one individual heart. Red lines represent loaded group, blue lines are unloaded group. Dotted line denoted 95% confidence band of the best-fit lines. P < 0.05 as statistic significant.

### Cell Cycling Markers Are Not Increased in Unloaded Hearts

To determine if decreased DNA in Unloaded hearts was associated with generation of new diploid cells by proliferation, we quantitated two markers of cell cycling: Ki67 and H3P. As a control, we measured Ki67-positivity in cultured cardiac fibroblasts, obtained from 4 separate subjects. In cultured fibroblasts, 17.4% ± 4.1% of cells were Ki67-positive (Figure 4A). We then quantitated Ki67 positivity in CM isolated from unloaded and loaded samples. Positive cells were rigorously defined by the presence of staining in the nucleus in combination with cytoplasmic α-actinin positivity (Figure 4B). Ki67+ CM were exceedingly rare over 5 loaded and 8 unloaded hearts; in total, 3 of 10579 and 7 of 54898 CM (from loaded and unloaded hearts, respectively) were positive for Ki67. Furthermore, only cells from 3 subjects (one loaded and two unloaded) showed any positivity (Figure 3C). Similar results were found using the G2/M marker H3P ^17^ in a smaller group of subjects (Supplemental Figure 3 and Supplemental Table 2).

**Figure 4.**
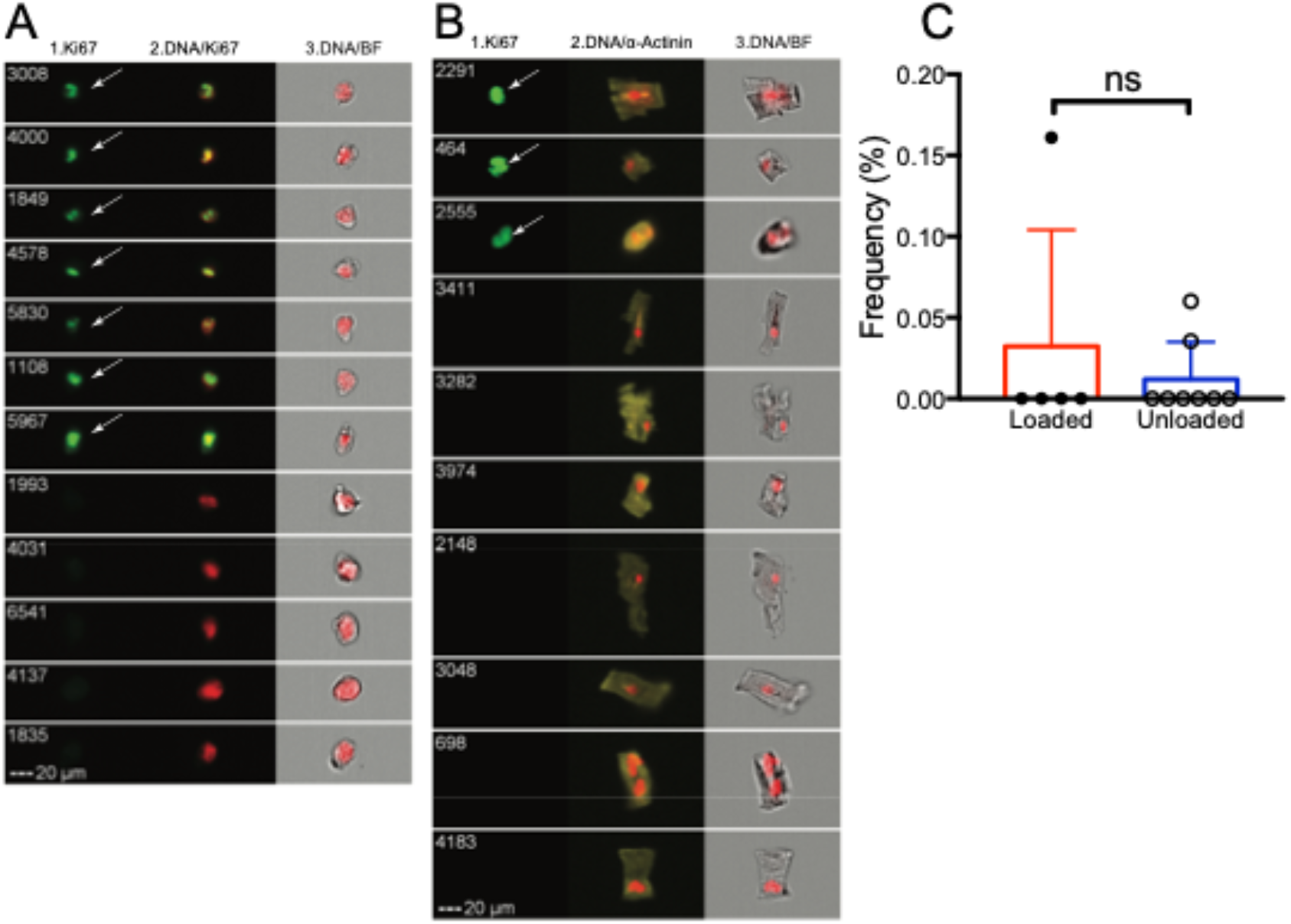
Cell cycling marker Ki67 does not increase with unloading. **A**, Representative images of cultured cardiac fibroblasts in the imaging flow cytometry analysis. **B**, Representative images of cardiomyocytes from an unloaded subject in the imaging flow cytometry analysis. White arrow denoted the Ki67 positive cells. **C**, Frequency of Ki67 positive cardiomyocytes. Each closed dot (loaded) or opened dot (unloaded) represent the mean value of one individual heart. Red bar represents loaded group, blue bar is unloaded group. Error bar represents standard deviation. Unpaired student t test was used to compare Ki67 between the groups. P < 0.05 as statistic significant. ns = not significant.

## DISCUSSION

Comparison of myocardium from failing hearts and failing hearts unloaded with LVAD support is a potent tool to uncover pathways to promote recovery of the failing heart ^13^. Multiple studies have shown that even at this late stage of disease, unloading of the heart leads to profound changes in myocardial cell structure and function. Reserve for regeneration perfect model to test. Using this model and a novel technique of imaging flow cytometry to unequivocally define cardiomyocytes, we were able to quantitate 10 times more cells per subject than previous studies ^15, 18-22^. This throughput allowed analysis of specific subpopulations of cardiomyocytes providing mechanistic insights into the reversal of hypertrophic remodeling that occurs with unloading.

With regard to morphologic changes with unloading, our data demonstrate cardiomyocytes from unloaded hearts are *<u>only mildly</u>* decreased in size compared to those from failing hearts. Although our data did not reach statistical significance, the 15% reduction in cardiomyocyte size was that consistent with three studies (19, 21, 23), that showed an average ∼20 percent decrease in size of CM. However, this finding is in contradistinction to other studies showing dramatic (nearly 50%) reductions in measured CM dimensions (15, 20, 22). It is possible that differences in measurement techniques account for the discrepancy between studies. In histologic analyses, CM are chosen for measurement by investigators based on typical morphology which introduces a human component that may exclude particularly large or small cardiomyocytes. Our unbiased method provided cardiomyocytes for analysis that were more variable in size than those measured in histologic studies. Furthermore, the percent decrease in CM size we measured was congruent with the difference in heart to body mass ratios between the Loaded and Unloaded groups. To our knowledge, 50% reductions in heart mass with LVAD unloading are uncommon.

More dramatic than changes in cell size were significant changes in cardiomyocyte DNA content. Whereas DNA content correlated with cell size in loaded hearts, with unloading DNA content dropped to a degree that this correlation was lost. Decreased cardiomyocyte DNA content following unloading has been previously reported and attributed to the generation of new diploid cells via proliferation^14, 15^. Different from those studies, our methodology allowed specific analysis of the both nucleation state and nuclear DNA content of intact CM. Our data demonstrate that there were no increases in the proportion of low DNA mononuclear cells in unloaded hearts–reduced DNA content was found in nuclei from both mono- and bi-nucleated CM. Furthermore, using imaging FACs to quantify cell cycle activity in cells that were unambiguously intact CM, we found only rare cells that were positive for Ki-67 or H3P with no differences in counts between loaded and unloaded hearts.

As both DNA content and cycling markers were measured at timepoints distant from LVAD implantation, these data do not rule out the possibility that a burst of proliferation immediately following LVAD implantation could account for changes in DNA content. Indeed, neither cell size nor DNA content changed with increasing time of LVAD unloading, implying that any changes occur acutely. On the other hand, mononuclear cell size was not disproportionally smaller in unloaded subjects. If new cells are generated in great numbers, it would be expected that this population would be distinctly smaller in size than existing cells due to lack of hypertrophic stimuli on LVAD support. Indeed, we saw important correlations between cell size, nuclear size and DNA content in failing hearts. Larger cells had more nuclei, but they were smaller with less DNA per nucleus than nuclei from mononuclear cells. These data suggest that mononucleated cardiomyocytes may develop a more robust cell cycling response to stress resulting in profound hyperploidy without cytokinesis. Further studies focused on this mononuclear population following LVAD implantation could not provide critical information on mechanisms that can stimulate cardiomyocyte regeneration.

If changes in DNA content are not due to new populations of cells, then what mechanism accounts for these changes? First, our observation could potentially be due to a loss of high ploidy cells in response to unloading via apoptosis or autophagy. Alternatively, the polyploidy that occurs in cardiomyocytes in the failing heart could regress in the absence of pressure overload. This process has been reported in animal models of hypertension in which aortic smooth muscle cell polyploidy decreases following treatment^23^. In other states, cells may increase copy numbers of genes that promote survival^24^. It is unknown if these increased copies regress when the state is removed. Finally, pieces of nucleus and DNA can undergo an autophagocytic process {nucleophagy ^25^} to decrease DNA content. Further studies using in situ hybridization techniques (outside the scope of this report) could help define these mechanisms.

Our study has several limitations. The cell isolation technique required more tissue that is available from an LVAD core so we could not perform studies on matched specimens from the same subject. As the end number of cells analyzed was far less than in the original tissue sample, cell isolation may select for a particular population of cells which may not represent what is occurring on the whole organ level. Moreover, cell isolation techniques may alter cell morphology and characteristics. However, all samples were treated identically and thus although absolute values may be skewed by the process, the differences between the groups is unlikely to be an artifact. Furthermore, Imagestream was able to measure CM of widely varying sizes (from ∼500 to 3500 um^2^) and CM size was dramatically different between normal size and function hearts and failing hypertrophied hearts, and our measures were on par with a previous study of dispersed CM pre- and post-LVAD unloading ^18^. Finally, our data could represent differential detection of the fluorescent output of the DNA bound dye due to changes in chromatin structure. Although these dyes bind stoichiometrically, it is unknown if dramatic changes in heterochromatin to euchromatin ratios could alter measurement of fluorescent output without changes in actual DNA content.

In conclusion, using a highly specific technique we have shown that demonstrate dynamic changes in cardiomyocyte DNA occur in multiple subsets of cells after hemodynamic unloading. These data add to this field by elucidating that the decrease in DNA content occurs independent of changes in cell size and is present in both mononuclear and binuclear cardiomyocytes. Furthermore, our data do not support the hypothesis that new cardiomyocytes are generated after LVAD unloading in the end-stage heart failure. However, they provide a roadmap to the isolation of specific cardiomyocyte populations and open up intriguing areas of research into mechanisms of changes in nuclear structure. Both will provide avenues to the discovery of novel strategies to induce myocardial recovery.

## Data Availability

All data is present in the paper

## Acknowledgements

The authors acknowledge Suzanne Steffes, RN for research coordination and Michelle Black and Christine Probst for the assistance with image flow cytometry analysis.

