## Supplemental Data for "Mechanical Unloading is Associated with Decreased DNA Content in Cardiomyocytes Independent of Nucleation State"

Supplemental Material includes:

**Supplemental Experimental Methods**

**Supplemental Figures and Legends**

**Supplemental Table and Legends**

**Supplemental References**

### Detailed Methods

#### Patient Clinical Characteristics

All patients had a complete clinical examination including echocardiography, right heart catheterization, and serum brain natriuretic peptide (BNP) measurement as part of routine clinical care. Data most proximal to the time of LVAD placement and/or cardiac transplantation was collected and analyzed. Echocardiograms were performed with standard techniques per clinical protocols. Left ventricular end-diastolic diameter (LVEDD), diastolic posterior wall thickness (PWd), and diastolic septal wall thickness (IVSd) were measured using Syngo Dynamics (Siemens Healthcare). LV mass indexed to body surface area ($g/m^2$) was calculated as described previously [1].

#### Specimen Procurement

The trimmed, wet weight of all explanted hearts were measured during pathologic examination. From each explanted heart, 5-10 strips from the mid-anterior wall of the left ventricle (epicardium to endocardium–approximately 500 mg, 5 x 5 x 20 $mm^3$ in size) were collected and stored in liquid nitrogen until analysis. An additional myocardial sample underwent immediate dissociation for fibroblast isolation. Specimen processing occurred within 1 hour of heart explant.

#### Dissociation of Cardiomyocytes

Myocyte isolation was done in batches. Briefly, frozen tissue was thawed in 4% paraformaldehyde (PFA, Sigma P6148) and cut into 1-2 $mm^3$ cubes. Fixation was achieved at room temperature in 4% PFA for 2 hours on a rotator followed by washing with PBS (HyClone SH30256.01) three times at room temperature. Fixed myocardial cubes were then transferred into conical tubes and incubated in PBS digest solution containing 1.8 mg/ml collagenase B (Roche, 11088823103), 2.4 mg/ml collagenase D

(Roche, 11088874103) and 75 µg /mL deoxyribonuclease-I (Sigma, D4513) overnight at 37°C as described previously[2].

**Isolation and Culture Expansion of Cardiac Fibroblasts**

Fresh cardiac samples were washed with warm Hank's balanced salt solution (ThemoFisher 14175-095) and minced in Iscove's modified Dulbecco's medium (IMDM, ThemoFisher 12440-053) supplemented with 1 mg/ml collagenase IV (ThemoFisher 17104-019), 0.15 mg/ml dispase (ThemoFisher 17105-041) and 75 µg/ml deoxyribonuclease-I (Sigma D4513). After incubating at 37 °C for 45 minutes, tissue was pelleted and triturated in 4°C IMDM followed by passage through a 40 µm cell strainer (Fisher Sci. 08-771-1). Filtered cells were washed and culture expanded at 37°C in the growth media (IMDM, 20% FBS, 100 U/ml penicillin G, 100 µg/ml streptomycin, 2 mmol/l L-glutamine, and 0.1 mmol/l 2-mercaptoethanol). Cells within 10 passages were harvested at 80 ~ 90% confluence then stored in PBS containing 1% paraformaldehyde. Cardiac fibroblasts were used within a week after harvesting and served as a control in parallel to each batched cardiomyocyte imaging flow cytometry analysis.

**Image Acquisition and Post-Acquisition Analysis**

To evaluate the nuclear makeups, cell sizes, DNA content and cell cycling events, cardiomyocyte images were acquired using INSPIRE software® on Amnis® imaging flow cytometer (ImageStream® Mark II, EMD Millipore). Data were acquired at x20 magnification from multiple 50 µL samples. The flow speed was approximately 60 ~ 132 mm/second. In the process of acquisitions, approximately 20mW of 488nm laser and 10mW of 642 nm laser were used to excite the fluorophores. High-resolution images of each cell were acquired directly from flow with the AF488 image in channel 2 (green), the AF555 images in channel 3 (orange), the AF700 images in channel 5 (far red) and the laser scatter image in channel 6. The brightfield images were acquired with 26 mW LED intensities on Channel 4. Because of the expected extremely low frequency of cell cycling events in the cardiomyocytes, which does not produce enough AF488 positive images in channel 4, cardiac fibroblasts were used to generate the matrix data for spectral compensation for each batched cardiomyocyte analysis. In this case, cell images of unstained and stained with single antibody and fluorophore combination were acquired without brightfield illumination.

Post-acquisition spectral compensation was performed using IDEAS® analysis software (EMD Millipore). To calculate the compensation matrix, single color fluorophore images were processed, the amount of spectral interfering between the three applied channels was determined by plotting intensity for the specific signal channel and the negative signal channel then a best-fit line was used to determine the amount of spectral overlap from each fluorescent marker in each channel. The adjusted values in the compensation matrix were then applied to each pixel of the cardiomyocyte image to create a data set that each signal sequestered into its intended spectral channel. The compensated cardiomyocyte images were validated by verifying that the median fluorescent intensity of the unlabeled cardiomyocytes in the overlapped channel was similar to the median fluorescent intensity of the single color positive control channel. Cardiomyocyte cell cycling frequency of positive image of Ki67 and H3P (**Supplemental Figure 4**) respectively. Briefly, images were first deconvolved to obtain in-focus, square images associated with fluorescent labels. For each event confirmed as an intact cell, multiple channel images were visualized simultaneously: brightfield, DNA content (Draq5), α-actinin (AF555), and cell cycle markers Ki67 and H3P (AF488) images were captured for analysis. Flow cytometry events determined to be cell fragments, or negative for α-actinin or DNA labels were excluded from analysis. Cardiomyocyte and nuclear size were measured using the brightfield and Draq5 staining images, respectively. Subfractions of mononuclear, binuclear, trinuclear (and beyond) cardiomyocytes were quantified by the spot count algorithm applied to a “spot mask” based on the objects positive to Draq5 staining.

These masking parameters were visually optimized for each sample. DNA content of each individual cardiomyocyte or of each individual nucleus was measured. Cell cycle activity was only confirmed when an appropriate marker (Ki67 or H3P) was present within the nuclear contour area in an intact cell (**Supplemental Figure 2**).

### Supplemental figure 1: ImageStream post-acquisition analysis

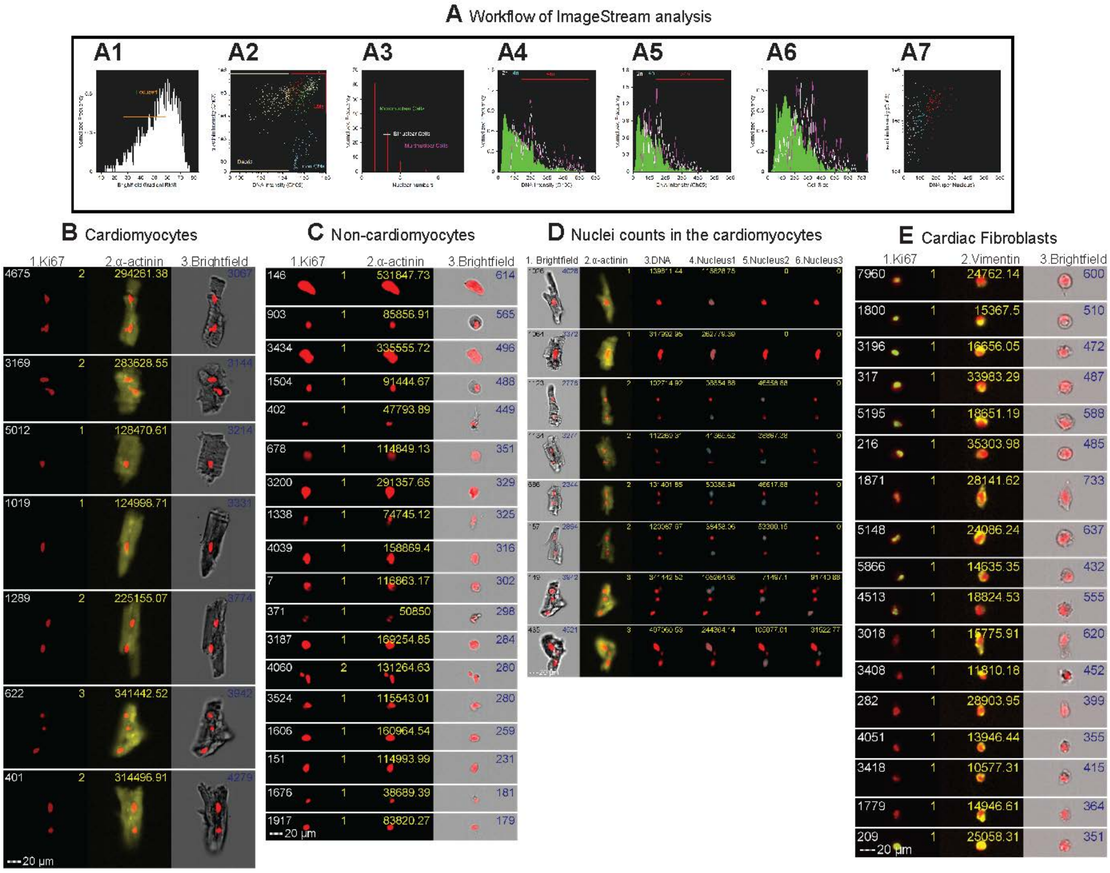


All the data files acquired from the same sample were combined in the post-acquisition analysis. Accumulatively 10,000 images were collected from each sample for cardiomyocyte morphologic profiling. Image analysis was completed using the algorithms integrated in the IDEAS® analysis software (6.2.65.0, EMD Millipore). Cells in best focus were identified using gradient RMS of the brightfield image (**Suppl.Fig2 A1**). This is followed by gating on cardiomyocytes (**Suppl.Fig2 A2**) which are the events that dual positive for α-actinin and DNA staining (**Suppl.Fig2 B**). Events negative to DNA staining are excluded as debris. Events of DNA positive only are consisted of a heterogenous particles, including cardiac fibroblasts and other non-cardiomyocytes (**Suppl.Fig2 C**). Next, to assess the nuclear numbers (**Suppl.Fig2 A3**), the cardiomyocyte images were analyzed by the algorithms for spot counting of puncta that are positive to Draq5 (**Suppl.Fig2 D**). Cardiomyocyte size (**Suppl.Fig2 A6**) was decided by the area measurement of brightfield images (**Suppl.Fig2 B-**

**column3 brightfield**), nuclear size was decided by the area of Draq5 staining (**Suppl.Fig2 D-column4-6**). DNA content was measured by quantifying the Draq5 fluorescent intensity of each cell (**Suppl.Fig2 D column3)** or the average intensity of each individual nucleus (**Suppl.Fig2 D column4-6**). Average cell size (**Suppl.Fig2 A6**) and nuclear size of sub-fractional population based on the nucleation state are calculated. There are more potential correlation analyses can be achieved such as cells with high DNA content correlated to higher α-actinin intensity (**Suppl.Fig2 A7**). In addition, in contrast to the rear frequency of Ki67 in the cardiomyocytes, positive Ki67 (Green) is 17.4% ± 4.1% in the cultured cardiac fibroblasts isolated from 4 subjects (**Suppl.Fig2 E**).

**Supplemental Figure 2. Heart mass does not correlate with cardiomyocyte size in either loaded or unloaded hearts.**

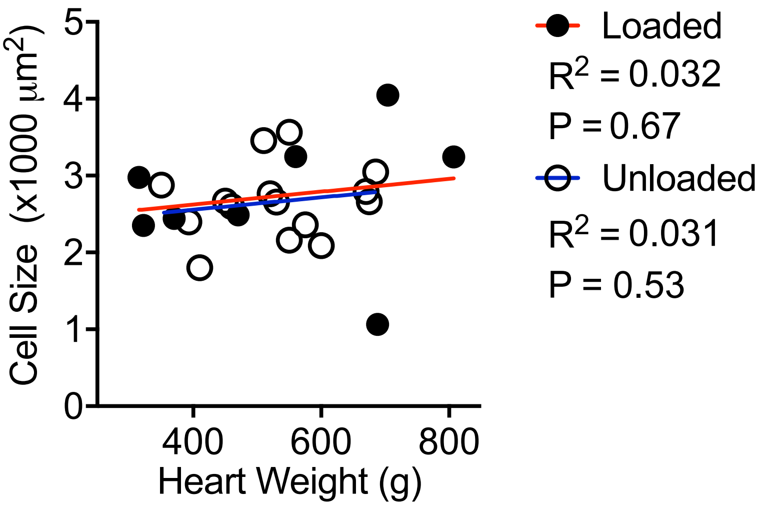


Simple linear regression analysis showed no correlation of cardiomyocyte size and heart mass in either loaded ($R^2$=0.03, P = 0.67) or unloaded heart ($R^2$=0.03, P = 0.53). Compare the overall slopes and the intercepts, there is no difference between loaded and unloaded hearts (the slopes P = 0.99 and the intercepts P = 0.80). Each closed dot (loaded) or opened dot (unloaded) represents the mean value of one individual heart. Red line represents loaded group; blue line is unloaded group. $P < 0.05$ as statistic significant.

**Supplemental Figure 3. Representative images of H3P positive and negative cardiomyocytes in the imaging cytometry**

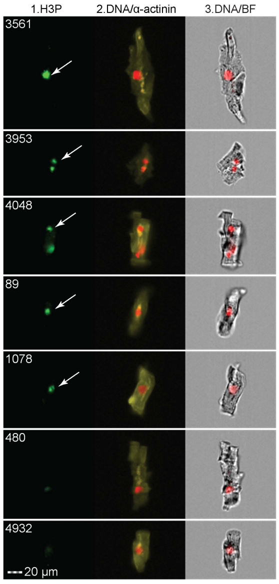


Cardiomyocyte nuclei that negative or positive (arrow) to H3P labeling (green) are showed in column1 (positive images are combined from 1 loaded and 3 unloaded samples). Nuclear images (red) are merged with positive α-actinin (yellow) or brightfield images and showed in column2 and 3 respectively. The white digits in column1 are the sequence numbers of the objects on the imaging cytometry. Scale bar = 20μm.

**Supplemental Table 1: General Patient Characteristics.**

| | CHF-Loaded | CHF-Unloaded | P Value |
|---|---|---|---|
| Total Enrolled (male:female) | 8 (5:3) | 15 (14:1) | |
| N = Ischemic cardiomyopathy | 3 | 4 | |
| Age (years) | 50.6 ± 14.7 | 58.4 ± 9.4 | 0.14 |
| Ejection Fraction (%) | 21.4 ± 4.4 | 17.5 ± 4.2 | 0.05 |
| Heart Weight (HW, g) | 529.5 ± 189.6 | 528.5 ± 103.4 | 0.99 |
| Body Weight (BW, kg) | 76 ± 18.7 | 92.4 ± 15.3 | 0.03 |
| HW-BW Ratio (g/kg) | 6.9 ± 1.5 | 5.8 ± 1.2 | 0.07 |
| Pre-LVAD BNP (μg/ml) | 1.1 ± 0.8 | 1.0 ± 0.5 | 0.81 |
| Post-LVAD BNP (μg/ml) | | 0.3 ± 0.3 | 0.03 |
| Duration of LVAD (months) | | 13.7 ± 9.1 | |

Data are presented as mean ± one standard deviation. Unpaired student t test was used to compare between the groups.

**Supplemental Table 2: Demonstration of remodeling in CHF-Unloaded subjects pre- and post-LVAD implantation**

| | Pre-LVAD | Post-LVAD | P Value |
|---|---|---|---|
| LVEDD (cm) | 7.7 ± 1 | 5.9 ± 0.9 | 7E-08 |
| LV Mass Index (g/m2) | 141.3 ± 33 | 106.2 ± 32.1 | 0.002 |
| BNP (μg/ml) | 0.99 ± 0.54 | 0.33 ± 0.27 | 0.001 |

Paired Student *t* test was applied for the comparison between pre- and post-LVAD in the unloaded group. Heart size and LV mass index were calculated based on the echocardiography.

**Supplemental Table 3: Quantification cardiomyocyte cell cycling events**

| | Ki67 | H3P |
|---|---|---|
| **Loaded** | | |
| Average of Number of CM studied | 2375 | 7971 |
| Number of hearts studied | 5 | 3 |
| Positive (%) | 0.032 | 0.005 |
| **Unloaded** | | |
| Average of Number of CM studied | 6612 | 2352 |
| Number of hearts studied | 8 | 6 |
| Positive (%) | 0.012 | 0.011 |

Comparison of the prevalence of positive proliferation markers in the cardiomyocytes, Unpaired Student *t* test with Bonferroni correction was used for the comparison between the two groups.
